# Naturalistic acceptance-based emotion regulation in adolescents with NSSI: altered prefrontal activation and amygdala–prefrontal connectivity

**DOI:** 10.64898/2026.05.03.26352312

**Authors:** Heng Jiang, Jingxian He, Liyuan Li, Yuxuan Guo, Xianyang Gan, Xiaoxia Fan, Xia Wang, Stefania Ferraro, Deniz Vatansever, Keith M Kendrick, Christian Keysers, Valeria Gazzola, Bo Zhou, Benjamin Becker

**Affiliations:** Sichuan Provincial Center for Mental Health, Sichuan Provincial People’s Hospital, School of Medicine, University of Electronic Science and Technology of China, Chengdu, China; Brain-Computer Interface & Brain-Inspired Intelligence Key Laboratory of Sichuan Province, School of Life Science and Technology, University of Electronic Science and Technology of China, Chengdu, China; Huangshui Primary School, Chengdu, China; Institute of Science and Technology for Brain-inspired Intelligence, Fudan University, Shanghai, China; Netherlands Institute for Neuroscience, KNAW, Amsterdam, The Netherlands; Department of Psychology, University of Amsterdam, Amsterdam, The Netherlands; MIND & AI Lab, Department of Psychology, The University of Hong Kong, Hong Kong SAR, China; Strategic Research Topic: AI, Society & Social Dynamics, Faculty of Social Sciences, The University of Hong Kong, Hong Kong SAR, China

**Author notes:** **Corresponding authors:** Bo Zhou, University of Electronic Science and Technology of China, Chengdu, China,; Benjamin Becker (lead contact), The University of Hong Kong, Hong Kong, China. **Abbreviations:** NSSI, non-suicidal self-injury; HC, health control; PHQ-9, Patient Health Questionnaire; DERS, Difficulties in Emotion Regulation Scale; ER, emotion regulation; NeutR, neutral video clips–react; NegR, negative video clips–react; NegA, negative video clips–acceptance; vlPFC, ventrolateral prefrontal cortex; OFC, orbitofrontal cortex; vmPFC, ventromedial prefrontal cortex; dlPFC, dorsolateral prefrontal cortex; dmPFC, dorsomedial prefrontal cortex; gPPI, generalized psychophysiological interaction; ROI, region of interest.

**Keywords:** non□suicidal self□injury (NSSI), emotion regulation, dynamic naturalistic emotional contexts, functional magnetic resonance imaging (fMRI)

## Abstract

**Background:** Non-suicidal self-injury (NSSI) represents a growing public health concern, particularly in adolescents. Emotion dysregulation is central to prevailing NSSI models, yet it remains unclear whether acceptance-based emotion regulation (ER) and its underlying neural processes are disrupted in naturalistic, dynamic contexts.

**Methods:** Pre-registered neuroimaging trial in recently diagnosed and treatment-naïve adolescents with NSSI (n=25) and healthy controls (n=25) using an ER paradigm with dynamic video clips and concomitant functional magnetic resonance imaging. Behavioral, neural activity, and connectivity indices during emotion reactivity and acceptance-based regulation were compared between groups.

**Results:** Adolescents with NSSI experienced elevated negative feelings during neutral clips, reflecting heightened baseline negativity. In comparison to controls, they displayed reduced temporal and ventrolateral prefrontal engagement during emotional reactivity, but increased engagement of regions implicated in both emotion reactivity (right amygdala, insula) and ER (right dlPFC, dmPFC, vlPFC) when utilizing acceptance. Higher activation in the right dlPFC was positively associated with difficulties in accessing ER strategies in everyday life. Adolescents with NSSI showed reduced functional connectivity between the right amygdala and left dlPFC.

**Conclusions:** Adolescents with NSSI exhibited a baseline negativity bias and altered neural engagement during both negative emotional reactivity and acceptance-based regulation, characterized by increased activation and reduced amygdala–dlPFC connectivity. These findings highlight atypical emotion processing in real-life contexts in individuals with NSSI. Targeting acceptance-based regulation and prefrontal–limbic circuitry may represent a promising intervention approach for adolescents with NSSI.

## Introduction

Non-suicidal self-injury (NSSI) is defined as the deliberate harm to one’s own body without suicidal intent and has been suggested as a psychiatric condition warranting further investigation in the Diagnostic and Statistical Manual of Mental Disorders, 5th edition (DSM-5) (American Psychiatric Association, 2013; Liu et al., 2016; Nock, 2010). NSSI in adolescents has emerged as a pressing public health concern with significant mental and physical health implications (Nock, 2010; Qu et al., 2023; Swannell et al., 2014; Wu et al., 2025). Recent epidemiological studies indicate high prevalence rates among adolescents, with lifetime and 12-month prevalence estimates of 20–50% and 20%, respectively (Qu et al., 2023; Wester et al., 2018; Xiao et al., 2022). NSSI also exhibits bidirectional relationships and high co-morbidity with multiple psychiatric disorders, in particular depression (Hasking et al., 2025; Wu et al., 2023), which poses a challenge for isolating the specific behavioral and neural pathways underlying the condition. Importantly, NSSI during adolescence serves as a predictor of psychiatric conditions in later adolescence and adulthood (Halpin & Duffy, 2020; Mars et al., 2014; Wilkinson et al., 2018).

Emotion dysregulation, a transdiagnostic construct across multiple mental disorders (Sloan et al., 2017), has consistently been identified as a core feature that contributes to the development and maintenance of NSSI (American Psychiatric Association, 2013; Hasking et al., 2017; Liu et al., 2016; Nock, 2009; Qu et al., 2023; Wolff et al., 2019). Effective emotion regulation (ER) requires adaptive selection and deployment of strategies to modulate emotional experiences (Gross & Hooria, 2014). However, individuals engaging in NSSI tend to rely on maladaptive strategies, such as rumination, catastrophizing, self-blame, and blaming others (Lang et al., 2024). A three-wave longitudinal study (∼two years) suggested a bidirectional relationship between emotion dysregulation and NSSI behavior, such that baseline emotion dysregulation predicted future NSSI engagement, and NSSI undermines ER abilities (Robinson et al., 2019). Neuroimaging studies in young adults further suggest that NSSI, during cognitive reappraisal, is associated with reduced recruitment of regulatory and emotional brain regions, including the superior temporal gyrus, putamen, thalamus, supplementary motor area, and inferior frontal gyrus (Kim et al., 2025). Given that adolescence is a period of ongoing neural and cognitive development, individuals in this stage are particularly vulnerable to emotional volatility and dysregulation (Byeon et al., 2026; He et al., 2025; Lee et al., 2014; Paus et al., 2008). The dynamic nature of real-life situations further exacerbates these challenges, as poor ER is associated with rigidity, heightened affective inertia, and difficulty flexibly adjusting emotional responses across contexts (Hollenstein, 2015).

Acceptance, a core component of third-wave mindfulness-based cognitive therapies, regulates negative affect via nonjudgmental awareness of emotional experiences, rather than attempting to control them (Bishop et al., 2004; Kober et al., 2019). Acceptance has been shown to facilitate the regulation of negative affect in both experimental and clinical settings while imposing relatively low cognitive demands (Goldin et al., 2019; Messina et al., 2021; Monachesi et al., 2023). However, individuals currently engaging in self-injury demonstrate significantly lower mindfulness than past self-injurers and non-self-injurers, and that low mindfulness significantly predicts engagement in self-injury (Caltabiano & Martin, 2017). Meta-analytic evidence also indicates that non-acceptance of emotional responses is strongly associated with NSSI behaviors (Wolff et al., 2019). Collectively, these findings suggest that adolescents with NSSI may show alterations in ER abilities when employing acceptance-based strategies. Yet, the neurobiological underpinnings of these potential deficits remain unclear, warranting further investigation to inform targeted behavioral, cognitive, and neural interventions.

Naturalistic ER paradigms, which utilize dynamic videos, speech, or music as stimuli, have emerged as an ecologically valid approach to assess ER in contexts that closely resemble real-life experiences (Jiang, He, Zimmermann, et al., 2026; Sonkusare et al., 2019; Zhou & Becker, 2025). In the current pre-registered study (ClinicalTrials.gov ID: NCT05907421), we applied a validated functional magnetic resonance imaging (fMRI) naturalistic ER paradigm (Jiang, He, Zimmermann, et al., 2026) using immersive video clips to examine acceptance-based regulation in adolescents with NSSI relative to healthy controls (HC). To address potential confounding effects of medication, we restricted recruitment to treatment-naïve adolescents with NSSI in the peak prevalence age range of 13-18 years (details see methods). We hypothesized that compared to HC, adolescents with NSSI would exhibit stronger negative emotional reactivity and show dysregulated neural recruitment of brain systems engaged in emotional reactivity and acceptance-based regulation (e.g., limbic regions, default mode network, and frontal regions). Neural activation was examined using whole-brain analyses, complemented by targeted seed-based functional connectivity analyses focusing on the right amygdala as a key region implicated in intense emotional reactivity and ER (Etkin et al., 2015; Morawetz et al., 2017; Ochsner et al., 2012; Zhang et al., 2025; Zimmermann et al., 2017).

## Methods

### Participants

A total of 30 recently diagnosed and treatment-naïve adolescents with NSSI behaviors were recruited from a local hospital, and 36 adolescents in the HC group were recruited via community advertisements. Exclusion criteria for both groups encompassed left-handedness, color vision deficiencies, smoking, and any current or past neurodevelopmental (e.g., Attention-Deficit Hyperactivity Disorder, ADHD) or neurological disorder, as well as any contraindications to MRI.

Participants in the NSSI group were clinically assessed according to DSM-5 criteria by an experienced clinician, and diagnoses were further confirmed using a validated self-report questionnaire (Buelens et al., 2020; Jiang, He, Zhou, et al., 2026). Additional inclusion criteria for the NSSI group were: (1) NSSI as the primary diagnosis, (2) NSSI being the main reason for seeking treatment, and (3) no prior behavioral or pharmacological interventions for psychiatric conditions, including NSSI. HC participants were required to have no current or past psychiatric disorders, including NSSI. Participants’ depressive symptoms were assessed using the Patient Health Questionnaire (PHQ-9, Kroenke et al., 2001), and their ER difficulties using the Difficulties in Emotion Regulation Scale (DERS, Gratz & Roemer, 2004).

Three NSSI participants did not complete the assessments; two NSSI and eight HC participants were excluded due to excessive head motion (> 6 mm or 6°); and three HC participants did not pass post-scan quality checks (Jiang, He, Zhou, et al., 2026; Jiang, He, Zimmermann, et al., 2026). Data were analyzed from 25 adolescents with NSSI and 25 HC participants who completed the full protocol and met all data-quality criteria.

All participants (and their legal guardians, if aged<18 years) provided written informed consent, and were compensated 100 RMB for participation. The present sample was drawn from a larger project on emotional dysregulations in NSSI, with prior findings on pain empathy already published (Jiang, He, Zhou, et al., 2026). The protocol for the present experiment was pre-registered. Of note, the pre-registered age range of 15–18 years was extended to 13–18 years to better match the peak prevalence rate of NSSI. The final sample encompassed 30 adolescents with NSSI and 36 HC, as opposed to the pre-registered 40 per group, due to recruitment challenges based on the strict inclusion criteria that focused on treatment-naïve adolescents with primary NSSI diagnosis.

All procedures were conducted in accordance with the Declaration of Helsinki, and the original and age-range extended trial was approved by the ethics committees of the University of Electronic Science and Technology of China and Sichuan Academy of Medical Sciences & Sichuan Provincial People’s Hospital.

### Stimuli and Paradigm

A validated naturalistic ER fMRI paradigm was employed to investigate the neural substrates of negative emotional reactivity and ER via acceptance (Fig. 1, Jiang, He, Zimmermann, et al., 2026, see Fig. 1). The stimuli used in the present study were adapted following consultations with professional clinicians to ensure age- and population-appropriate content (e.g., avoiding depictions of physical aggression). Stimuli were validated in an independent sample of adolescents (n=20), who rated the clips on valence and arousal. In addition, temporal variability in valence and arousal within each clip was assessed to ensure consistency of emotional experiences. The paradigm contained two types of stimuli: neutral and negative. Three conditions were used: NeutR (neutral-react) and NegR (negative-react), in which participants were instructed to respond naturally to the stimuli without attempting to regulate or reduce their emotional reactions to neutral or negative clips; and NegA (negative-acceptance), in which participants were instructed to experience their emotions within an “inaction” framework, without judging, controlling, changing, or resisting their feelings. After each clip, participants rated their momentary negative affect on a 9-point Likert scale (1-not negative at all, 9-very negative); at the end of each run, they rated their average success level (1-not successful at all, 9-very successful). The selected clips under NegR and NegA were matched in terms of valence, arousal, and consistency. Written instructions were provided, along with standardized verbal explanations delivered by a single experimenter. Participants completed a practice session before the experiment, which included four scanning runs of nine trials each, with three trials per condition randomly presented.

**Fig.1.**
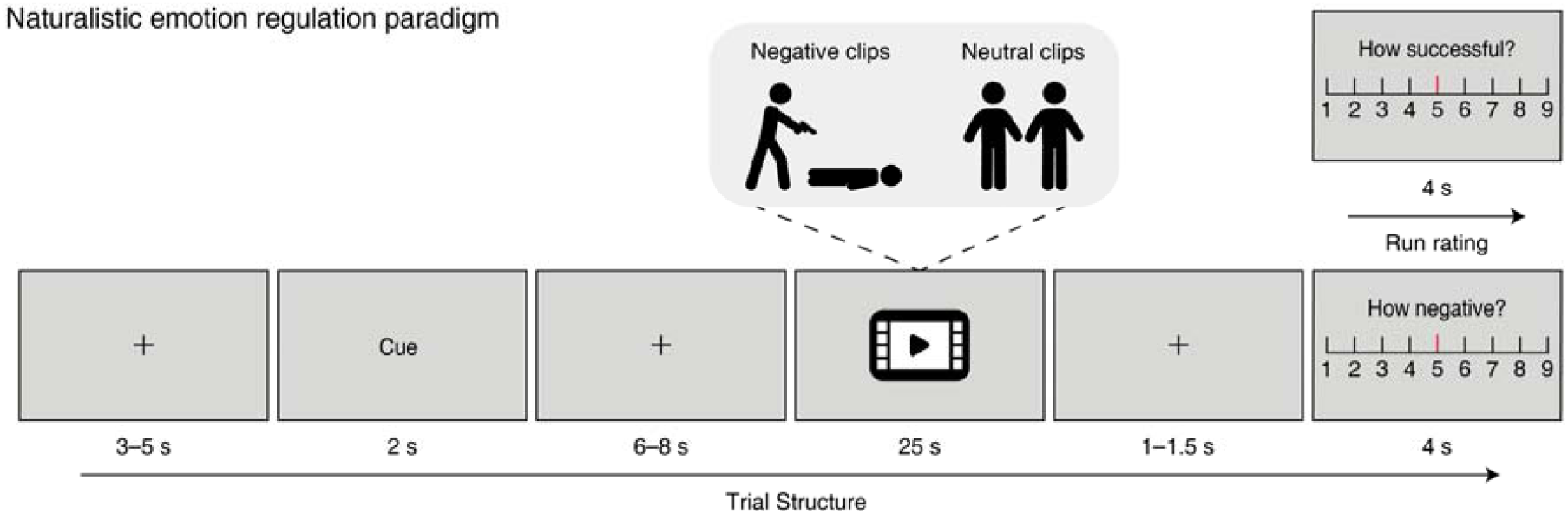
Illustration of the paradigm structure. The naturalistic emotion regulation (ER) paradigm is used to measure emotion reactivity and ER ability. The cues were “react” or “acceptance”, indicating which strategy should be used to view the neutral or negative video clips. Neutral clips were always paired with the “react” cue (NeutR), whereas negative clips were paired with either the “react” (NegR) or “acceptance” (NegA) cue, with 12 trials per condition. The video clips were matched across conditions in terms of contextual features (e.g., activities and social content). In addition, no significant differences were observed between NegR and NegA in valence, arousal, or within-clip temporal variability. Trials were presented in a random order. At the end of each trial, participants rated their negative feelings (1-not negative at all, 9-very negative), and at the end of each run, they rated their average success level (1-not successful at all, 9-very successful). Adapted from Jiang, He, Zimmermann, et al. (2026). Notably, the schematics were included only to circumvent copyright restrictions and do not represent the original stimuli.

### Behavioral analyses

Averaged participant-level negative affect ratings were analyzed using separate 2 (stimulus type: NeutR, NegR)×2 (group: HC, NSSI) and 2 (strategy: NegR, NegA)×2 (group: HC, NSSI) mixed ANOVAs. End-of-run success ratings were compared using independent-sample t-test.

### MRI data acquisition and preprocessing

MRI data were acquired on a 3.0T Siemens Vida scanner with a 64-channel head coil. High-resolution structural images were obtained and used for structural localization and transformation into Montreal Neurological Institute (MNI) space. Functional data were collected with an echo planar imaging–free induction decay sequence. Preprocessing was conducted using Statistical Parametric Mapping (SPM12). The first five functional volumes were discarded, followed by slice timing correction, realignment with unwarping, co-registration, segmentation, normalization to MNI space (2×2×2 mm), spatial smoothing with an 8-mm FWHM Gaussian kernel, and bias-field correction. Motion-related outliers were identified using standardized criteria and included as nuisance regressors (Gan et al., 2024; Jiang, He, Zhou, et al., 2026; Jiang, He, Zimmermann, et al., 2026). The mean number of outlier volumes did not differ between groups (HC: 12.41±19.13, M±SD; NSSI: 11.83±15.90; *t*=0.117, *p*=0.908). Further acquisition and preprocessing details are provided in the Supplementary Information.

### Mass-univariate analyses and ROI-based Exploration

Subject-level general linear model (GLM) analysis was conducted using SPM12. Nuisance regressors included 24 head motion parameters and outlier time points (Gan et al., 2024; Jiang, He, Zhou, et al., 2026). The fixation-cross periods served as the implicit baseline. Clip presentation served as the main regressor of interest, while cue and rating periods were modeled as regressors of no interest. A high-pass filter of 180s was applied. To account for individual differences in behavioral responses, the subjective negativity ratings for each clip were included as a parametric modulator, consistent with our previous study (Jiang, He, Zimmermann, et al., 2026). Key contrasts were NegR>NeutR (emotional reactivity) and NegA>NegR (acceptance-based regulation. To control for the potential confounding effect due to group differences in depressive symptom severity, individual PHQ-9 scores were included as a covariate in the second-level group analyses (Aybek et al., 2015). Statistical maps were thresholded using a cluster-level Family-Wise Error (FWE) correction of (*p*<0.05) with a cluster-forming threshold of *p*<0.001_uncorrected_.

To characterize large-scale neural engagement during acceptance, the spatial similarity between contrast maps and seven large-scale resting-state networks (Yeo et al., 2011), as well as prefrontal systems commonly involved in ER (dlPFC, dorsolateral prefrontal cortex; vlPFC, ventrolateral prefrontal cortex; dmPFC, dorsomedial prefrontal cortex; vmPFC, ventromedial prefrontal cortex; OFC, orbitofrontal cortex; ACC, anterior cingulate cortex; Carlén, 2017), was illustrated by river plots (Gan et al., 2024; Jiang, He, Zimmermann, et al., 2026). The spatial similarity was computed as cosine similarity between the networks or regions of interest (ROIs) and the beta map of NegA>NegR for each group separately, reflecting the extent to which the NegA>NegR activation pattern was expressed within each predefined region.

Given prior meta-analytic evidence linking ER difficulties to NSSI (Wolff et al., 2019), we conducted regression analyses to examine their association with brain activation during acceptance and ER difficulties. To identify brain regions engaged in acceptance processing across both groups for subsequent correlational analyses with ER implementation in everyday life, while avoiding circular analysis (Kriegeskorte et al., 2009), data from adolescents with NSSI and HC were pooled, and NegA vs. NegR contrast was computed. One cluster in the right prefrontal region showing increased activation was defined as ROI, encompassing premotor regions and dlPFC, within a premotor–prefrontal transitional zone. While dlPFC subregions are implicated in cognitive control and ER (Dixon et al., 2017; Etkin et al., 2015; Morawetz et al., 2017; Ochsner et al., 2012), the broader anatomical extent suggests additional premotor contributions. A small portion of this cluster overlapped with regions showing group differences during acceptance. Mean Blood-Oxygen-Level Dependent (BOLD) signal within the ROI was extracted using the MarsBar toolbox (version 0.45; Brett et al., 2002). Analyses were restricted to two theoretically and empirically relevant DERS subscales: Limited Access to Emotion Regulation Strategies and Nonacceptance of Emotional Responses, which have shown robust and consistent associations with NSSI in prior meta-analytic work (Wolff et al., 2019). Group and depression level measured by PHQ-9 were included as a covariate. All *p*-values within the current analyses were False Discovery Rate (FDR) corrected.

### Connectivity analyses

Functional connectivity was assessed using the generalized psychophysiological interaction (gPPI) approach implemented in CONN (version v.22.v2407; Whitfield-Gabrieli & Nieto-Castanon, 2012), which estimates task-dependent functional connectivity by modeling condition-specific interactions between a seed region and whole-brain activity through deconvolved and reconvolved BOLD time series (McLaren et al., 2012). Based on previous studies demonstrating an important role of the right amygdala in intense emotional experiences and of right amygdala-frontal connectivity in ER deficits (Etkin et al., 2015; Morawetz et al., 2017; Ochsner et al., 2012; Zhang et al., 2025; Zimmermann et al., 2017), we examined between-group differences in seed-to-whole brain voxel-wide connectivity of this region. The 6-mm radius spherical seed (peak MNI: 20, −8, −22), encompassing the right amygdala extending into the hippocampus, was defined based on our prior findings using this naturalistic ER task in an independent sample of young adults employing the acceptance strategy (Jiang, He, Zimmermann, et al., 2026). This right-lateralized definition was further supported by the current results, only the right amygdala shown significant group difference.

On the subject-level analysis, regressors included the canonical hemodynamic responses for each psychological condition, the seed region time series, their interaction, as well as nuisance covariates (head motion parameters, outlier time points, white matter and cerebrospinal fluid signals, and psychological regressors). The residual BOLD time series were band-pass filtered (0.008=Hz to Inf). On the group-level analysis, we compared acceptance-related connectivity differences (NegA□>□NegR) between the NSSI and HC groups, while controlling for their depression level. Statistical maps were thresholded at voxel-level *p*□<□0.001_uncorrected_ and cluster-level *p*□<□0.05 (cluster-size FDR-corrected).

## Results

### Demographic and clinical characteristics

The demographic and clinical characteristics are presented in Table 1. The independent-samples t-test found that adolescents with NSSI exhibited significantly higher depression and ER difficulties levels compared to the HC.

**Table 1.** Demographics and clinical characteristics of adolescents with NSSI and HC participants.

|  | HC (n=25) |  | NSSI (n=25) |  | Statistical analysis |  |  |
| --- | --- | --- | --- | --- | --- | --- | --- |
|  | Mean | SD | Mean | SD | T | p-value | Effect sizes |
| **Demographic data** |  |  |  |  |  |  |  |
| Age (years) | 14.48 | 1.90 | 14.64 | 1.52 | -0.329 | 0.744 | 1.75 |
| Sex (male: female ([male])) | 8:17 | 32% | 13:12 | 52% | 2.052 ( $\chi^2$ ) | 0.15 | 0.20 |
| Educational level (years) | 9.08 | 1.82 | 9.04 | 1.40 | 0.087 | 0.931 | 1.65 |
| **Clinical data** |  |  |  |  |  |  |  |
| PHQ-9 | 4.80 | 4.48 | 19.88 | 4.39 | -12.018 | <.001 | 4.51 |
| DERS Non-acceptance | 10.40 | 4.31 | 21.00 | 5.94 | -7.224 | <.001 | 5.27 |
| DERS Goals | 12.20 | 4.90 | 19.52 | 4.26 | -5.636 | <.001 | 4.67 |
| DERS Impulse | 5.84 | 4.80 | 15.44 | 5.42 | -6.631 | <.001 | 5.20 |
| DERS Awareness | 16.00 | 5.13 | 20.72 | 4.77 | -3.370 | <.001 | 5.03 |
| DERS Strategies | 14.72 | 6.11 | 30.84 | 5.62 | -9.702 | <.001 | 5.97 |
| DERS Clarity | 7.00 | 3.04 | 9.36 | 3.01 | -2.757 | <.001 | 3.08 |
| DERS Total | 66.16 | 21.81 | 116.88 | 20.49 | -8.473 | <.001 | 21.50 |

PHQ-9, Patient Health Questionnaire (PHQ-9); DERS, Difficulties in Emotion
Regulation Scale; Non-acceptance, Nonacceptance of Emotional Responses; Goals,
Difficulties Engaging in Goal-Directed Behavior; Impulse, Impulse Control
Difficulties; Awareness, Lack of Emotional Awareness; Strategies, Limited Access to
Emotion Regulation Strategies; Clarity, Lack of Emotional Clarity.

### Subjective experience and regulation of negative emotions

A 2 (stimulus type: NeutR, NegR)×2 (group: HC, NSSI) mixed analysis of variance (ANOVA) revealed a significant main effect of stimulus type (*F*[1,48]=243.251, *p*=2.01×10^-20^, 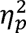 =0.835, see Fig. 2a). Negative clips induced more negative emotion (5.6±0.25) than the neutral (2.0±0.13). The interaction effect was significant (*F*[1,48]=4.765, *p*=3.4×10□², 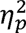 =0.090). Further simple effects analysis indicated that compared to the HC (1.51±0.19), the adolescents with NSSI (2.47±0.19) had significantly higher negative emotion induced by the neutral clips. No significant main effect of group was observed (*P*=0.166).

**Fig.2.**
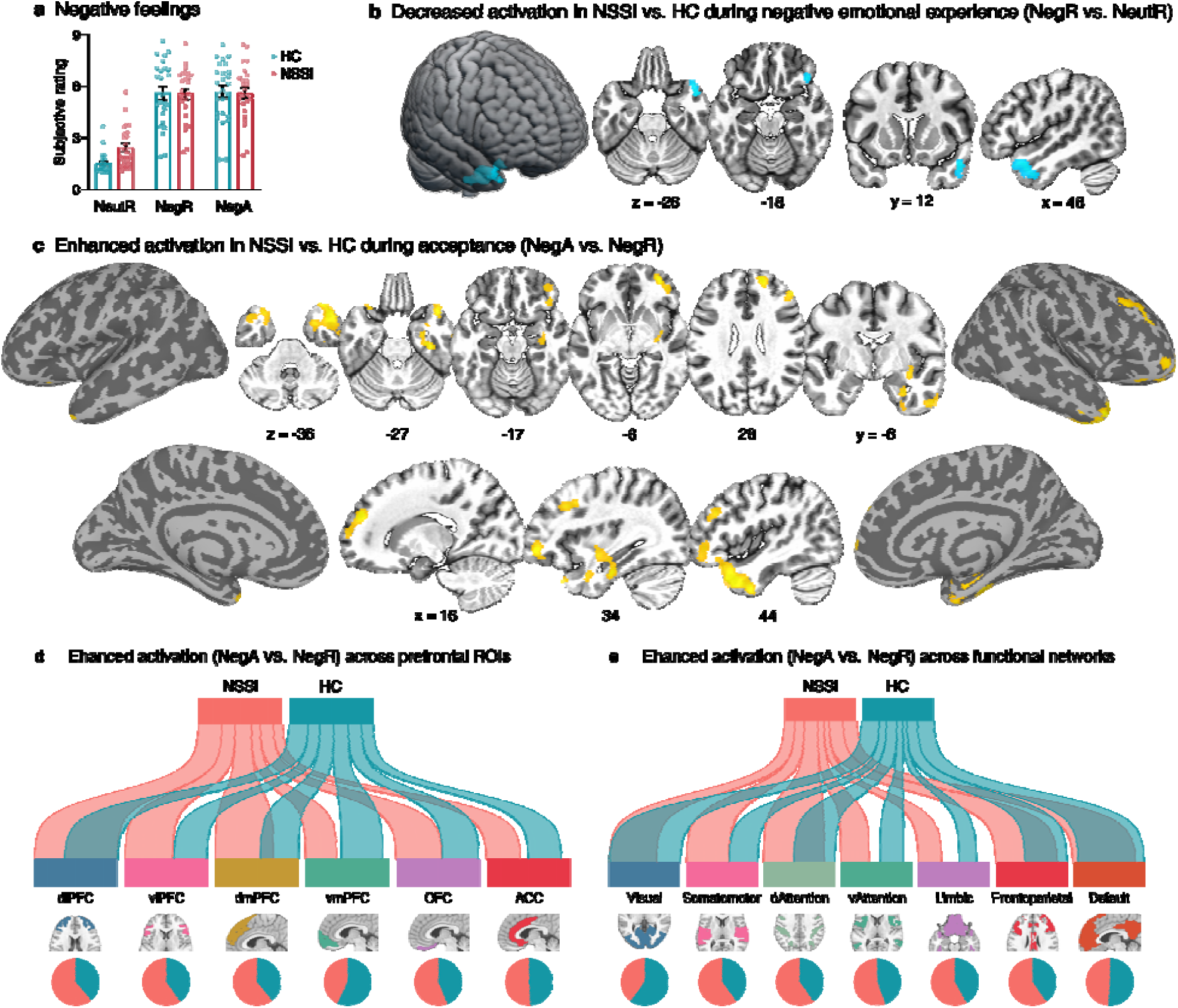
Subjective negative emotional state and brain activations. **a**, Average subjective feeling ratings of the adolescents with NSSI and the HC participants, during reacting to neutral (NeutR), negative (NegR) clips, and while applying the acceptance strategy to negative (NegA) video clips. Each dot represents the negative rating for individual participants averaged across trials within each condition. **b**, Brain regions showing greater activation in HC adolescents compared with those with NSSI, during naturalistic negative emotional reactivity (NegR>NeutR). **c**, Brain regions showing greater activation in adolescents with NSSI compared with HC adolescents when using the acceptance strategy to regulate negative emotions (NegA>NegR). Statistical maps are thresholded at voxel-level *p*<0.001 and cluster-level FWE-corrected at *p*<0.05. River plots showing spatial similarity (cosine similarity) between acceptance-related brain activation (NSSI and healthy controls analyzed separately) and (**d**) anatomically defined prefrontal cortex subdivisions (Carlén, 2017), or (**e**) resting-state-based functional cortical networks (Yeo et al., 2011). Ribbon thickness represents normalized maximum cosine similarity. Pie charts indicate the relative contribution of each activation pattern to each ROI (**d**) or network (**e**), defined as the proportion of voxels showing the highest cosine similarity within that region. dlPFC, dorsolateral prefrontal cortex; vlPFC, ventrolateral prefrontal cortex; dmPFC, dorsomedial prefrontal cortex; vmPFC, ventromedial prefrontal cortex; OFC, orbitofrontal cortex; ACC, anterior cingulate cortex; dAttention, dorsal attention; vAttention, ventral attention.

A separate 2 (strategy: NegR, NegA)×2 (group: HC, NSSI) mixed ANOVA examining ER revealed no significant main effects or interaction on negative affective ratings. Complementary results from a Bayesian ANOVA are shown in Table S1. Examining the self-experienced success ratings revealed that both groups reported relatively high levels of success, with no significant group difference (HC: 7.25±1.16, NSSI: 6.95±1.20, *p*=0.379).

### Adolescents with NSSI show decreased brain activation during negative emotional reactivity

During naturalistic negative emotional reactivity (NegR vs. NeutR), compared to HC, adolescents with NSSI showed decreased activation in the right anterior temporal cortex, mainly encompassing the temporal pole, anterior superior temporal gyrus, with smaller extensions into the right orbitofrontal and ventrolateral prefrontal cortex (OFC and vlPFC, see Fig. 2b and Table 2). The results without subjective negativity ratings as parametric modulator remain consistent (Fig.S1a). To control for higher negative emotion in adolescents with NSSI under the NeutR condition, group activation differences were examined separately for NegR and NeutR, showing no effects in NeutR and consistent reduced activation in NSSI during NegR (Fig.S1b), indicating that the effects were driven by group differences related to reacting to negative emotional clips.

**Table 2.**
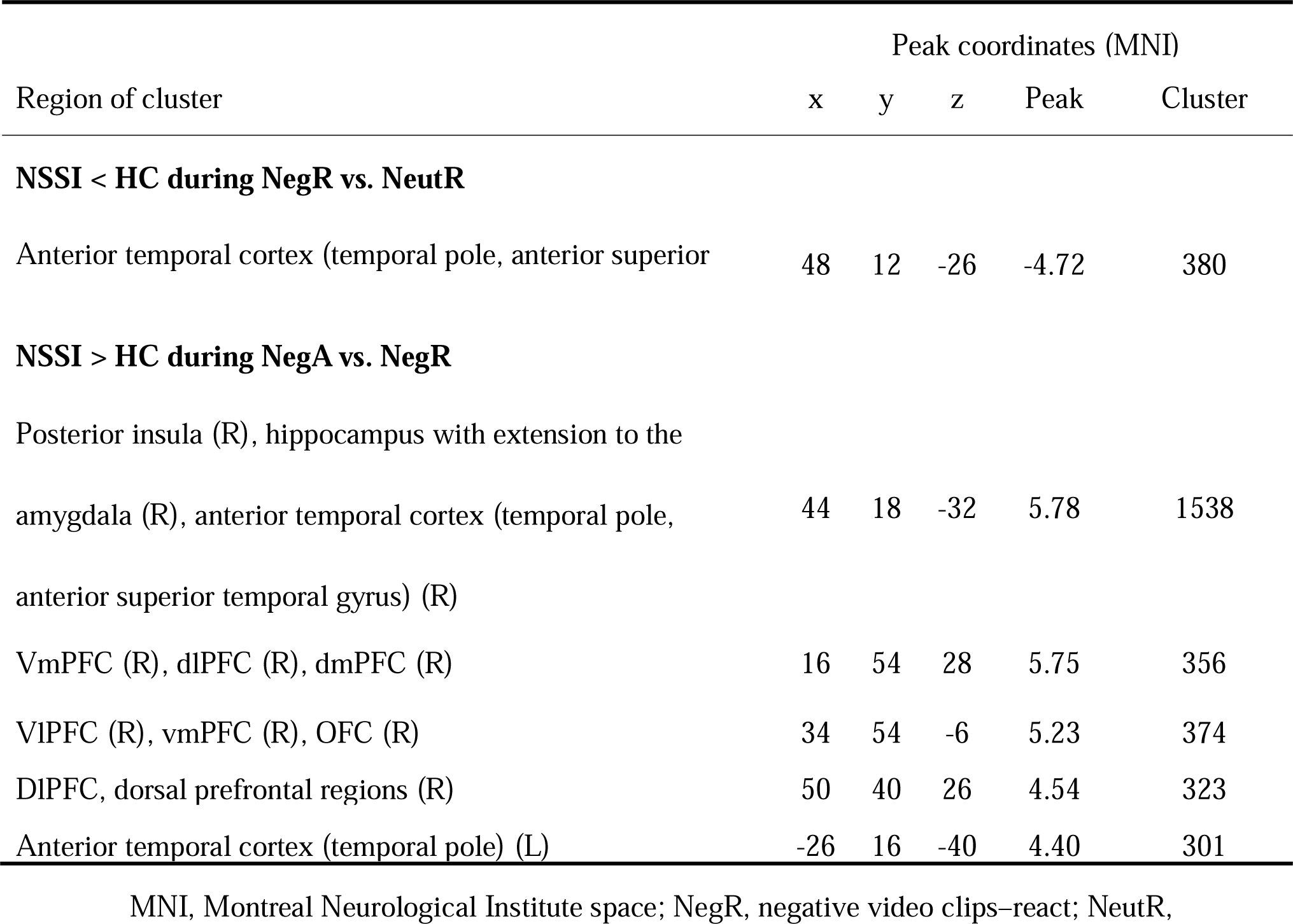
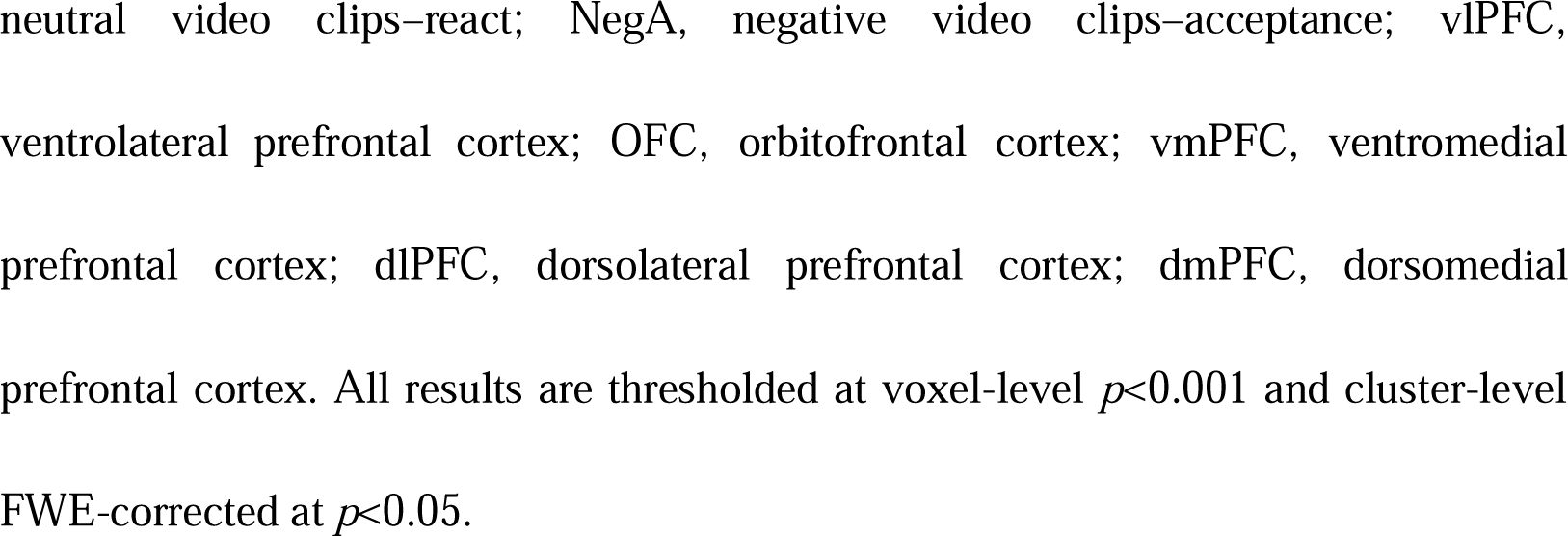
Regions show differential activations.

### Adolescents with NSSI show increased activity during acceptance

Compared with the HC participants, adolescents with NSSI exhibited significantly greater activation during acceptance (NegA vs. NegR) in regions encompassing higher-order cognitive control and ER, including the right dorsolateral and ventrolateral prefrontal cortices (dlPFC, vlPFC), as well as regions implicated in emotional processing, such as the right insula and hippocampus, with extension to the amygdala (see Fig. 2c and Table 2). Additionally, increased activation was observed in the bilateral anterior temporal cortex, encompassing the temporal pole and adjacent anterior temporal regions.

Pattern similarity analyses revealed that adolescents with NSSI showed greater similarity to reference patterns within multiple prefrontal ROIs that play crucial roles in ER, including dlPFC, vlPFC, dmPFC, and OFC (Fig. 2d). At the network level, NSSI participants’ activation regions were more similar to canonical networks, particularly the frontoparietal control, limbic, dorsal, and ventral attention, as well as somatomotor networks, indicating broader recruitment of distributed functional networks. These findings further suggest broader differences in the spatial organization of acceptance-related activation in adolescents with NSSI, in addition to the group differences in regional activation magnitude.

### Exploratory analyses: associations between brain activation during acceptance and emotion regulation difficulties

We further explored the association between neural responses and ER difficulties. DERS subscale scores and mean activation extracted from a significant cluster (including right premotor regions and dlPFC) identified in the NegA>NegR contrast (across groups) were entered into a regression model, controlling for group and depression severity. Results showed that higher activation was significantly positively associated with higher scores on Limited Access to Emotion Regulation Strategies (β=0.016, FDR *q*=0.034), but not with Nonacceptance of Emotional Responses (FDR *q*=0.315; see Fig. 3).

**Fig.3.**
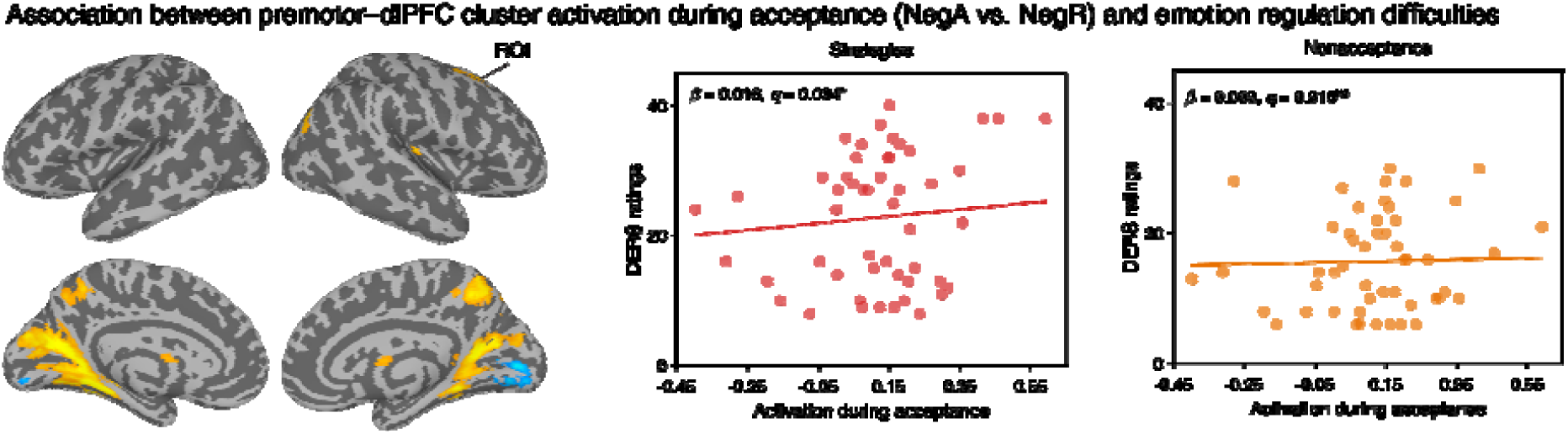
Associations of emotion regulation difficulties with brain activation as well as functional connectivity during acceptance. Left: The cortical map illustrates brain regions engaged during acceptance (NegA–NegR) across both groups combined. The significant cluster, including right premotor regions and dorsolateral prefrontal cortex (dlPFC), a region critically involved in ER, was defined as the region of interest (ROI). Right: Scatterplots show FDR-corrected associations between mean activation within the posterior dlPFC and scores on the Difficulties in Emotion Regulation Scale (DERS), specifically Limited Access to Emotion Regulation Strategies (Strategies) and Nonacceptance of Emotional Responses (Nonacceptance). Group and depression severity were included in the regression model as covariates of no interest. Dots represent individual participants, and shaded areas indicate the 95% confidence interval of the regression line. All results were FDR-corrected. NS, not significant; *, *p*<0.005.

### Adolescents with NSSI exhibit altered functional connectivity during acceptance

Seed-to-voxel connectivity analyses revealed that, during acceptance (NegA vs. NegR), compared with the HC participants, adolescents with NSSI showed significantly reduced functional connectivity between the right amygdala (with extension into the hippocampus) and the left dlPFC (peak MNI: −28,50,38; size 143, cluster size *p*-FWE-corrected=0.0410; see Fig. 4).

**Fig.4.**
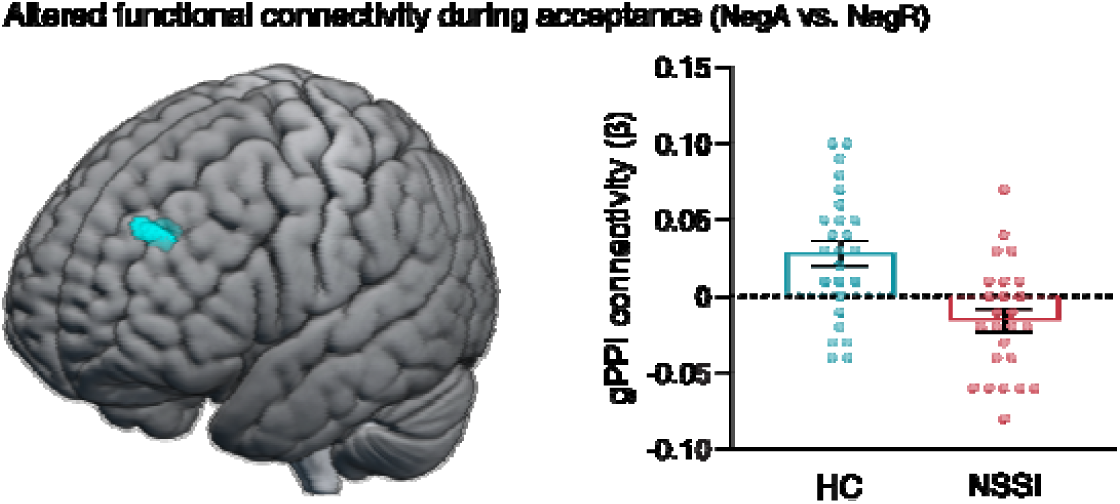
Reduced functional connectivity in adolescents with NSSI during acceptance. Left: Three-dimensional renderings showing that the left dlPFC exhibited significantly reduced functional connectivity with the right amygdala (seed) in adolescents with NSSI compared with healthy controls. Results were thresholded at voxel-level *p*<0.001 (uncorrected) and cluster-level *p*<0.05 (FDR-corrected). Right: gPPI connectivity (β) within the significant cluster in healthy controls and adolescents with NSSI.

## Discussion

As an increasingly prevalent public health concern with profound mental and physical health consequences, particularly during adolescence, NSSI has been robustly linked to deficits in emotion-related processes (American Psychiatric Association, 2013; Hasking et al., 2017; Liu et al., 2016; Nock, 2009; Qu et al., 2023; Wolff et al., 2019). To our knowledge, the present study is the first neurofunctional investigation of acceptance-based ER in adolescents with NSSI using a close-to-real-life naturalistic paradigm. Consistent with our broad hypotheses, adolescents with NSSI reported greater negative affect during neutral clips and showed altered activation in the negative-reactivity and instructed-acceptance contrasts, including reduced right temporal/OFC/vlPFC responses during NegR>NeutR and increased prefrontal and insula/medial-temporal responses during NegA>NegR.

Beyond regulation itself, our findings raise the possibility that adolescents with NSSI show altered context-sensitive emotional processing, reflected here in elevated negative affect during neutral scenarios. Adolescents with NSSI reported elevated negative affect during neutral scenarios, indicating atypical emotional reactivity. While no significant group differences in brain activation were observed during reacting naturally to neutral video alone (NeutR), isolating individuals’ negative emotional reactivity while controlling for responses to neutral videos (NegR-NeutR) revealed that adolescents with NSSI exhibited reduced brain activations in the temporal cortex, vlPFC, and OFC—regions that have been implicated in updating and contextual modulation of emotional information (Ochsner et al., 2012; Rolls et al., 2020; Silvers et al., 2015). Moreover, similar patterns were observed when examining the NegR condition alone (see Fig.S1), suggesting that the altered emotional reactivity in adolescents with NSSI was primarily related to responses to negative emotional scenarios. Consistent with this interpretation, prior work has reported heightened facial electromyographic reactivity to both positive and negative stimuli in adolescents with NSSI despite null group differences in fMRI responses and self-report measures (Mayo et al., 2021). In young adults, NSSI has likewise been associated with altered neural responses to negative stimuli, including aberrant activation in the amygdala, anterior cingulate cortex, orbitofrontal cortex, and temporal regions (particularly the right superior temporal gyrus), as well as the parahippocampal and supramarginal gyri (Hooley et al., 2020; Kim et al., 2025). Together with previous studies and theoretical models of NSSI, the present findings are compatible with the view that altered emotional reactivity and context-sensitive emotional processing are important components of the clinical phenotype (Chapman et al., 2006; Nock, 2009). Importantly, these alterations may manifest not as exaggerated responses to overtly negative stimuli per se, but rather as context-insensitive or poorly modulated emotional processing. Future studies with larger samples spanning a broader range of ages and NSSI severity are needed to further clarify these mechanisms.

A central finding was that adolescents with NSSI showed atypical neurofunctional engagement during the instructed acceptance of negative emotions. Acceptance is generally conceptualized as a relatively low cognitive load ER strategy that emphasizes self-referential and metacognitive awareness (Goldin et al., 2019). In line with our recent work in young adults using a similar naturalistic regulation paradigm (with some variation in negative stimulus content; Jiang, He, Zimmermann, et al., 2026), adolescents with NSSI demonstrated enhanced activation in brain regions implicated in interoceptive processing (posterior insula; Craig, 2011; J. Li et al., 2019),stimulus valuation (vmPFC; Ochsner et al., 2012), and subcortical emotional reactivity (hippocampus with extension to the amygdala; Bo et al., 2024; Doré et al., 2018; Kober et al., 2019) during acceptance implementation. Moreover, they exhibited hyperactivation in multiple prefrontal regions associated with high-level cognitive control and ER, including the dorsolateral and ventrolateral prefrontal cortices (Etkin et al., 2015; Morawetz et al., 2017; Ochsner et al., 2012). Additionally, temporal regions (including the temporal pole), as well as the vlPFC and OFC, showed increased activation during acceptance-based ER, and these regions have been implicated in updating and contextual modulation of emotional information (Ochsner et al., 2012; Rolls et al., 2020; Silvers et al., 2015). Their enhanced activation may be compatible with greater recruitment of regulatory or control-related processes during instructed acceptance. Beyond these regional effects, spatial similarity analyses further indicated that task-evoked activation patterns in NSSI were more extensively aligned with multiple prefrontal regions (dlPFC, vlPFC, dmPFC, OFC) and large-scale functional networks (somatomotor, attention, limbic, and frontoparietal systems). These findings suggest that the regional activation differences occurred within a broader spatial pattern involving prefrontal and large-scale functional networks, although this analysis should be interpreted descriptively. Importantly, this distributed organization does not preclude region-specific functional relevance. Activation whitin a right premotor–prefrontal cluster, encompassing premotor and dlPFC commonly implicated in cognitive control processes during ER (Etkin et al., 2015; Morawetz et al., 2017; Ochsner et al., 2012), was positively associated with greater difficulties in accessing ER strategies. However, given that this cluster extends into posterior premotor cortex and that the present task involved dynamic, action-rich video clips, this activation may also partly reflect altered processing of observed actions or embodied resonance during movie viewing, rather than exclusively canonical dlPFC-based ER (Gazzola, Rizzolatti, et al., 2007; Gazzola, Worp, et al., 2007; Gazzola & Keysers, 2009; Jimenez et al., 2020). Notably, functional connectivity analyses further revealed reduced coupling between the right amygdala, a region implicated in emotional reactivity, and the left dlPFC, indicating less efficient integration between prefrontal control and limbic reactivity in adolescents with NSSI. This pattern is consistent with prior evidence demonstrating that prefrontal–limbic integration plays a crucial role in successful ER (Banks et al., 2007; Berboth & Morawetz, 2021). Importantly, although both groups reported relatively high levels of perceived regulation success during acceptance (mean ratings ∼7 on a 9-point scale), this subjective experience was not accompanied by a robust reduction in negative affect at the behavioral level. This dissociation suggests that participants may have engaged with the instructed strategy without achieving measurable changes in emotional output at the behavioral level. Taken together, these findings suggest that adolescents with NSSI may recruit prefrontal and limbic systems differently during instructed acceptance, a pattern that could reflect greater regulatory demands or altered coordination between affective and control-related systems.

Comparable patterns of inefficient cognitive recruitment during ER have been documented across multiple clinical populations. Specifically, during the most commonly studied reappraisal strategy, individuals with depression (Johnstone et al., 2007), panic disorder (Reinecke et al., 2015), bipolar disorder (Morris et al., 2012), and drug abuse (Zimmermann et al., 2017, 2018) commonly exhibit exaggerated prefrontal activation, which has been interpreted as compensatory recruitment of cognitive resources. Neuroimaging evidence of acceptance-based strategies in individuals with emotional alterations is more limited but similarly indicates altered neural responses. For example, individuals with remitted major depressive disorder exhibit reduced activation in the right frontal pole when using acceptance to regulate picture-induced negative emotions (Smoski et al., 2015), while a region-of-interest analysis found lower bilateral insula activation during acceptance in patients with borderline personality disorder (Fernando et al., 2023). In the current study, adolescents with NSSI showed increased activation in the right insula during acceptance-based ER. This pattern may reflect disorder-specific differences or the possibility that dynamic naturalistic stimuli require greater cognitive and interoceptive engagement to implement acceptance. Altered dlPFC–amygdala connectivity during ER using reappraisal has been reported in several psychiatric conditions, including anorexia nervosa (Steward et al., 2022), major depression (Erk et al., 2010), and bipolar disorder (Zhang et al., 2018), highlighting disrupted integration between regions involved in emotional reactivity and higher-order control. Consistent with these findings, meta-analytic evidence indicates that neural differences in cognitive ER may emerge even when self-reported regulatory success does not differ between groups (Zilverstand et al., 2017). Analogously, in the present study, despite no significant group differences in self-reported effectiveness of acceptance-based regulation during fMRI, both groups reported relatively high levels of success. In contrast, adolescents with NSSI exhibited distinct neural activation patterns and reduced amygdala–dlPFC connectivity, each showing differential associations with ER difficulties, suggesting increased cognitive effort and less efficient prefrontal–limbic integration.

Emotion dysregulation has long been proposed as a central mechanism in the development and maintenance of NSSI (American Psychiatric Association, 2013; Chapman et al., 2006; Hasking et al., 2017; Liu et al., 2016; Nock, 2009; Qu et al., 2023; Wolff et al., 2019). Adolescents who lack effective regulatory strategies may resort to self-injury as a means of alleviating intense negative affect (Chapman et al., 2006; Hasking et al., 2017; Wang et al., 2024). The Experiential Avoidance Model posits that self-harm temporarily reduces aversive emotional states, thereby reinforcing NSSI as a maladaptive coping mechanism (Chapman et al., 2006). Similarly, Nock’s integrative model conceptualizes NSSI as a behavior maintained by its immediate regulatory benefits, arising from distal vulnerabilities such as heightened emotional reactivity and further shaped by learning processes (Nock, 2009). The ongoing maturation of prefrontal–limbic circuitry during adolescence may further exacerbate susceptibility to emotional instability (Lee et al., 2014; Paus et al., 2008). Together, the present findings provide neurofunctional support for these theoretical frameworks and deepen our understanding of emotion dysregulation in adolescent NSSI.

Acceptance has been proposed as an ER strategy that requires relatively fewer cognitive resources (Goldin et al., 2019). Given evidence that adolescents with NSSI may exhibit deficits in cognitive resource allocation (Fikke et al., 2011; Li et al., 2025), acceptance may represent a particularly suitable intervention target. Although the present findings suggest that acceptance is more effortful for adolescents with NSSI than for HC, systematic reviews indicate that with practice, acceptance-based regulation may increasingly rely on bottom–up processes (Chiesa et al., 2013). Supporting this, a recent study demonstrated that a 3-week mindfulness-based intervention significantly reduced negative emotions in adolescents with NSSI during an ER task, with effects maintained at a two-month follow-up (Zheng et al., 2024), highlighting the potential efficacy of acceptance-focused interventions. Future studies should leverage neuroimaging techniques to examine interventions aimed at strengthening acceptance and enhancing the ER efficiency in adolescents with NSSI, as well as their underlying neural mechanisms. Promising approaches include acceptance-and mindfulness-based therapies (Hofmann & Asmundson, 2008), and, potentially, adjunctive noninvasive neuromodulation techniques such as transcranial magnetic stimulation (Ding et al., 2025; Wang et al., 2025) and neurofeedback (Tsang et al., 2025). The prefrontal regions identified in the current study (e.g., right dlPFC) serve as potential neural targets, given the established role of dlPFC in ER models (Dixon et al., 2017; Etkin et al., 2015; Ochsner et al., 2012), and consistent evidence implicating amygdala–prefrontal circuitry in emotional regulatory control processes (Morawetz et al., 2025; Steward et al., 2022), although the present findings should be interpreted cautiously regarding specificity and directionality.

Several limitations warrant consideration. First, the cross-sectional design precludes causal inferences regarding whether the observed neural differences represent predisposing vulnerabilities or consequences of repeated self-injury. Second, although depression was statistically controlled, adolescents with NSSI reported elevated depressive symptoms, consistent with their high comorbidity (Marshall et al., 2013; Perini et al., 2019; Quevedo et al., 2016); future studies should include clinical comparison groups matched on depressive severity but without NSSI to better isolate NSSI-specific effects. Third, despite relatively high self-reported success rates, acceptance did not significantly reduce negative emotions. Future studies may benefit from examining multi-stage training protocols with varying durations to further validate the present findings. Fourth, the ROI-based brain–behavior analyses were exploratory, as the ROI was defined based on the functional contrast across groups, and not independently; accordingly, these findings should be interpreted as hypothesis-generating. Finally, the present findings should be interpreted in light of the sample characteristics, and future studies are needed to replicate and extend these results in larger and more diverse cohorts.

### Conclusion

Adolescents engaging in NSSI exhibited atypical neural responses during both negative emotional reactivity and acceptance-based ER. Hyperactivation during acceptance, together with its association with ER difficulties, suggests increased cognitive demands when implementing this strategy. Reduced amygdala–dlPFC connectivity further points to incomplete functional integration during regulation and was associated with limited access to ER strategies. Interventions targeting acceptance-based regulation and prefrontal–limbic integration may hold promise for improving ER in adolescents with NSSI.

## Data Availability

Data availability is not allowed due to institutional and ethical restrictions.

## Acknowledgements

This work is supported by the National Natural Science Foundation of China (NSFC 82271583, 82401812), the Ministry of Science and Technology of China (STI 2030–Major Projects 2022ZD0208500), the Hong Kong University Grants Council (GRF 17615525), the University of Hong Kong seed funding and start-up schemes (2407102536; 2402101713), the Health Commission of Sichuan Province Medical Science and Technology Program (24LCYJPT18), and the China Scholarship Council Program (No. 202506070094). Funders were not involved in the research design, the collection and analysis of data, the decision to publish, or the writing of the manuscript.

## References

American Psychiatric Association. (2013). Diagnostic and statistical manual of mental disorders (Fifth Edition). Arlington, VA, American Psychiatric Association.

Aybek, S., Nicholson, T. R., O’Daly, O., Zelaya, F., Kanaan, R. A., & David, A. S. (2015). Emotion-Motion Interactions in Conversion Disorder: An fMRI Study. PLOS ONE, 10(4), e0123273. 10.1371/journal.pone.0123273

Banks, S. J., Eddy, K. T., Angstadt, M., Nathan, P. J., & Phan, K. L. (2007). Amygdala–frontal connectivity during emotion regulation. Social Cognitive and Affective Neuroscience, 2(4), 303–312. 10.1093/scan/nsm029

Berboth, S., & Morawetz, C. (2021). Amygdala-prefrontal connectivity during emotion regulation: A meta-analysis of psychophysiological interactions. Neuropsychologia, 153, 107767. 10.1016/j.neuropsychologia.2021.107767

Bishop, S. R., Lau, M., Shapiro, S., Carlson, L., Anderson, N. D., Carmody, J., Segal, Z. V., Abbey, S., Speca, M., Velting, D., & Devins, G. (2004). Mindfulness: A proposed operational definition. Clinical Psychology: Science and Practice, 11(3), 230–241. 10.1093/clipsy.bph077

Bo, K., Kraynak, T. E., Kwon, M., Sun, M., Gianaros, P. J., & Wager, T. D. (2024). A systems identification approach using Bayes factors to deconstruct the brain bases of emotion regulation. Nature Neuroscience. 10.1038/s41593-024-01605-7

Brett, M., Anton, J.-L., Valabregue, R., & Poline, J.-B. (2002). Region of interest analysis using the MarsBar toolbox for SPM 99. Neuroimage, 16(2), S497.

Buelens, T., Luyckx, K., Kiekens, G., Gandhi, A., Muehlenkamp, J. J., & Claes, L. (2020). Investigating the DSM-5 criteria for non-suicidal self-injury disorder in a community sample of adolescents. Journal of Affective Disorders, 260, 314–322. 10.1016/j.jad.2019.09.009

Byeon, K., Park, H., Park, S., Cluce, J., Mehta, K., Cieslak, M., Cui, Z., Hong, S.-J., Chang, C., Smallwood, J., Satterthwaite, T. D., Milham, M. P., & Xu, T. (2026). Developmental variations in recurrent spatiotemporal brain propagations from childhood to adulthood. Nature Communications. 10.1038/s41467-025-67754-w

Caltabiano, G., & Martin, G. (2017). Mindless Suffering: The Relationship Between Mindfulness and Non-Suicidal Self-Injury. Mindfulness, 8(3), 788–796. 10.1007/s12671-016-0657-y

Carlén, M. (2017). What constitutes the prefrontal cortex? Science, 358(6362), 478–482.

Chapman, A. L., Gratz, K. L., & Brown, M. Z. (2006). Solving the puzzle of deliberate self-harm: The experiential avoidance model. Behaviour Research and Therapy, 44(3), 371–394. 10.1016/j.brat.2005.03.005

Chiesa, A., Serretti, A., & Jakobsen, J. C. (2013). Mindfulness: Top–down or bottom–up emotion regulation strategy? Clinical Psychology Review, 33(1), 82–96. 10.1016/j.cpr.2012.10.006

Craig, A. D. (2011). Significance of the insula for the evolution of human awareness of feelings from the body. Annals of the New York Academy of Sciences, 1225(1), 72–82. 10.1111/j.1749-6632.2011.05990.x

Ding, X., Jiang, H., Kang, T., Wang, Y., He, L., Xie, R., Chu, X., Yin, Q., & Becker, B. (2025). Modulation of Emotion Regulation Deficits in Patients with Heroin Use Disorder by Intermittent Theta-Burst Transcranial Magnetic Stimulation. Research Square. 10.21203/rs.3.rs-6301058/v1

Dixon, M. L., Thiruchselvam, R., Todd, R., & Christoff, K. (2017). Emotion and the prefrontal cortex: An integrative review. Psychological Bulletin, 143(10), 1033–1081. 10.1037/bul0000096

Doré, B. P., Rodrik, O., Boccagno, C., Hubbard, A., Weber, J., Stanley, B., Oquendo, M. A., Miller, J. M., Sublette, M. E., Mann, J. J., & Ochsner, K. N. (2018). Negative Autobiographical Memory in Depression Reflects Elevated Amygdala-Hippocampal Reactivity and Hippocampally Associated Emotion Regulation. Biological Psychiatry: Cognitive Neuroscience and Neuroimaging, 3(4), 358–366. 10.1016/j.bpsc.2018.01.002

Erk, S., Mikschl, A., Stier, S., Ciaramidaro, A., Gapp, V., Weber, B., & Walter, H. (2010). Acute and Sustained Effects of Cognitive Emotion Regulation in Major Depression. Journal of Neuroscience, 30(47), 15726–15734. 10.1523/JNEUROSCI.1856-10.2010

Etkin, A., Büchel, C., & Gross, J. J. (2015). The neural bases of emotion regulation. Nature Reviews Neuroscience, 16(11), Article 11. 10.1038/nrn4044

Fernando, S. C., Beblo, T., Lamers, A., Schlosser, N., Woermann, F. G., Driessen, M., & Toepper, M. (2023). Neural correlates of emotion acceptance and suppression in borderline personality disorder. Frontiers in Psychiatry, 13, 1066218. 10.3389/fpsyt.2022.1066218

Fikke, L. T., Melinder, A., & Landrø, N. I. (2011). Executive functions are impaired in adolescents engaging in non-suicidal self-injury. Psychological Medicine, 41(3), 601–610. 10.1017/S0033291710001030

Gan, X., Zhou, F., Xu, T., Liu, X., Zhang, R., Zheng, Z., Yang, X., Zhou, X., Yu, F., Li, J., Cui, R., Wang, L., Yuan, J., Yao, D., & Becker, B. (2024). A neurofunctional signature of subjective disgust generalizes to oral distaste and socio-moral contexts. Nature Human Behaviour. 10.1038/s41562-024-01868-x

Gazzola, V., & Keysers, C. (2009). The Observation and Execution of Actions Share Motor and Somatosensory Voxels in all Tested Subjects: Single-Subject Analyses of Unsmoothed fMRI Data. Cerebral Cortex, 19(6), 1239–1255. 10.1093/cercor/bhn181

Gazzola, V., Rizzolatti, G., Wicker, B., & Keysers, C. (2007). The anthropomorphic brain: The mirror neuron system responds to human and robotic actions. Neuroimage, 35(4), 1674–1684.

Gazzola, V., Worp, H. van der, Mulder, T., Wicker, B., Rizzolatti, G., & Keysers, C. (2007). Aplasics Born without Hands Mirror the Goal of Hand Actions with Their Feet. Current Biology, 17(14), 1235–1240. 10.1016/j.cub.2007.06.045

Goldin, P. R., Moodie, C. A., & Gross, J. J. (2019). Acceptance versus reappraisal: Behavioral, autonomic, and neural effects. Cognitive, Affective, & Behavioral Neuroscience, 19(4), 927–944. 10.3758/s13415-019-00690-7

Gratz, K. L., & Roemer, L. (2004). Multidimensional Assessment of Emotion Regulation and Dysregulation: Development, Factor Structure, and Initial Validation of the Difficulties in Emotion Regulation Scale. Journal of Psychopathology and Behavioral Assessment, 26(1), 41–54. 10.1023/B:JOBA.0000007455.08539.94

Gross, J. J., & Hooria, J. (2014). Emotion, Emotion Regulation, and Psychopathology: An Affective Science Perspective. Clinical Psychological Science, 2, 387. 10.1177/2167702614536164

Halpin, S. A., & Duffy, N. M. (2020). Predictors of non-suicidal self-injury cessation in adults who self-injured during adolescence. Journal of Affective Disorders Reports, 1, 100017. 10.1016/j.jadr.2020.100017

Hasking, P., Kiekens, G., Petukhova, M. V., Albor, Y., Al-Hadi, A., Alonso, J., Al-Saud, N., Altwaijri, Y., Andersson, C., Atwoli, L., Muaka, C. A., Báez-Mansur, P., Ballester, L., Bantjes, J., Baumeister, H., Bendtsen, M., Benjet, C., Berman, A., Bruffaerts, R.,…Collaborators, W. M. H.-I. C. S. (2025). The relationships between sporadic and repetitive non-suicidal self-injury and mental disorders among first-year college students: Results from the World Mental Health International College Student Initiative. Psychological Medicine, 55, e280. 10.1017/S0033291725100688

Hasking, P., Whitlock, J., Voon, D., & Rose, A. (2017). A cognitive-emotional model of NSSI: Using emotion regulation and cognitive processes to explain why people self-injure. Cognition and Emotion, 31(8), 1543–1556. 10.1080/02699931.2016.1241219

He, S., Li, C., Zhang, S., Hu, Y., Wen, J., Liu, L., Gao, J., Wu, J., & Huang, G. (2025). The complexity of associations between emotion regulation, interpersonal sensitivity, cognitive insight, and non-suicidal self-injury: A study based on network analysis. BMC Psychiatry, 25(1), 846. 10.1186/s12888-025-07231-2

Hofmann, S. G., & Asmundson, G. J. G. (2008). Acceptance and mindfulness-based therapy: New wave or old hat? Clinical Psychology Review, 28(1), 1–16. 10.1016/j.cpr.2007.09.003

Hollenstein, T. (2015). This Time, It’s Real: Affective Flexibility, Time Scales, Feedback Loops, and the Regulation of Emotion. Emotion Review, 7(4), 308–315. 10.1177/1754073915590621

Hooley, J. M., Dahlgren, M. K., Best, S. G., Gonenc, A., & Gruber, S. A. (2020). Decreased Amygdalar Activation to NSSI-Stimuli in People Who Engage in NSSI: A Neuroimaging Pilot Study. Frontiers in Psychiatry, 11. 10.3389/fpsyt.2020.00238

Jiang, H., He, J., Zhou, B., Guo, Y., Gan, X., Fan, X., Wang, X., Ferraro, S., Vatansever, D., Kendrick, K. M., Li, L., & Becker, B. (2026). Adolescents with non-suicidal self-injury exhibit increased pain empathic neural reactivity and personal distress to physical but not affective pain. Journal of Affective Disorders, 399, 121145. 10.1016/j.jad.2025.121145

Jiang, H., He, J., Zimmermann, K., Zhou, X., Gan, X., Ferraro, S., Wang, L., Zhou, B., Li, L., Kendrick, K. M., Zhao, W., Yao, D., Yuan, T., Zhou, F., & Becker, B. (2026). Common and distinct neurofunctional signatures of dynamic naturalistic emotion regulation strategies. Nature Communications. 10.1038/s41467-026-70708-5

Jimenez, K. B., Abdelgabar, A.-R., De Angelis, L., McKay, L. S., Keysers, C., & Gazzola, V. (2020). Changes in brain activity following the voluntary control of empathy. NeuroImage, 216, 116529.

Johnstone, T., Reekum, C. M. van, Urry, H. L., Kalin, N. H., & Davidson, R. J. (2007). Failure to Regulate: Counterproductive Recruitment of Top-Down Prefrontal-Subcortical Circuitry in Major Depression. Journal of Neuroscience, 27(33), 8877–8884. 10.1523/JNEUROSCI.2063-07.2007

Kim, G., Shin, H., & Hur, J.-W. (2025). Differential neural activity associated with emotion reactivity and regulation in young adults with non-suicidal self-injury. BJPsych Open, 11(5), e163. 10.1192/bjo.2025.10765

Kober, H., Buhle, J., Weber, J., Ochsner, K. N., & Wager, T. D. (2019). Let it be: Mindful acceptance down-regulates pain and negative emotion. Social Cognitive and Affective Neuroscience, 14(11), 1147–1158. 10.1093/scan/nsz104

Kriegeskorte, N., Simmons, W. K., Bellgowan, P. S. F., & Baker, C. I. (2009). Circular analysis in systems neuroscience: The dangers of double dipping. Nature Neuroscience, 12(5), 535–540. 10.1038/nn.2303

Kroenke, K., Spitzer, R. L., & Williams, J. B. W. (2001). The PHQ-9: Validity of a brief depression severity measure. Journal of General Internal Medicine, 16(9), 606–613. 10.1046/j.1525-1497.2001.016009606.x

Lang, A. N., Zhong, Y., Lei, W., Xiao, Y., Hang, Y., Xie, Y., Lv, Z., Zhang, Y., Liu, X., Liang, M., Zhang, C., Zhang, P., Yang, H., Wu, Y., Wang, Q., Yang, K., Long, J., Liu, Y., Wang, S.,…Wang, C. (2024). Neural mechanism of non-adaptive cognitive emotion regulation in patients with non-suicidal self-injury. Comprehensive Psychiatry, 133, 152487. 10.1016/j.comppsych.2024.152487

Lee, F. S., Heimer, H., Giedd, J. N., Lein, E. S., Šestan, N., Weinberger, D. R., & Casey, B. J. (2014). Adolescent mental health—Opportunity and obligation. Science, 346(6209), 547–549. 10.1126/science.1260497

Li, J., Xu, L., Zheng, X., Fu, M., Zhou, F., Xu, X., Ma, X., Li, K., Kendrick, K. M., & Becker, B. (2019). Common and Dissociable Contributions of Alexithymia and Autism to Domain-Specific Interoceptive Dysregulations: A Dimensional Neuroimaging Approach. Psychotherapy and Psychosomatics, 88(3), 187–189. 10.1159/000495122

Li, S., Tian, X., Zhang, Y., Yang, Y., Sun, H., Chen, J., Xu, D., Xu, F., Zhu, X., Wang, K., & Qiao, D. (2025). Non-suicidal self-injury links with multidimensional cognitive dysfunctions in adolescents with depression. BMC Psychiatry. 10.1186/s12888-025-07701-7

Liu, R. T., Cheek, S. M., & Nestor, B. A. (2016). Non-suicidal self-injury and life stress: A systematic meta-analysis and theoretical elaboration. Clinical Psychology Review, 47, 1–14. 10.1016/j.cpr.2016.05.005

Mars, B., Heron, J., Crane, C., Hawton, K., Lewis, G., Macleod, J., Tilling, K., & Gunnell, D. (2014). Clinical and social outcomes of adolescent self harm: Population based birth cohort study. BMJ, 349, g5954. 10.1136/bmj.g5954

Marshall, S. K., Tilton-Weaver, L. C., & Stattin, H. (2013). Non-Suicidal Self-Injury and Depressive Symptoms During Middle Adolescence: A Longitudinal Analysis. Journal of Youth and Adolescence, 42(8), 1234–1242. 10.1007/s10964-013-9919-3

Mayo, L. M., Perini, I., Gustafsson, P. A., Hamilton, J. P., Kämpe, R., Heilig, M., & Zetterqvist, M. (2021). Psychophysiological and Neural Support for Enhanced Emotional Reactivity in Female Adolescents With Nonsuicidal Self-injury. Biological Psychiatry: Cognitive Neuroscience and Neuroimaging, 6(7), 682–691. 10.1016/j.bpsc.2020.11.004

McLaren, D. G., Ries, M. L., Xu, G., & Johnson, S. C. (2012). A generalized form of context-dependent psychophysiological interactions (gPPI): A comparison to standard approaches. NeuroImage, 61(4), 1277–1286. 10.1016/j.neuroimage.2012.03.068

Messina, I., Grecucci, A., & Viviani, R. (2021). Neurobiological models of emotion regulation: A meta-analysis of neuroimaging studies of acceptance as an emotion regulation strategy. Social Cognitive and Affective Neuroscience, 16(3), 257–267. 10.1093/scan/nsab007

Monachesi, B., Grecucci, A., Ahmadi Ghomroudi, P., & Messina, I. (2023). Comparing reappraisal and acceptance strategies to understand the neural architecture of emotion regulation: A meta-analytic approach. Frontiers in Psychology, 14, 1187092. 10.3389/fpsyg.2023.1187092

Morawetz, C., Bode, S., Derntl, B., & Heekeren, H. R. (2017). The effect of strategies, goals and stimulus material on the neural mechanisms of emotion regulation: A meta-analysis of fMRI studies. Neuroscience & Biobehavioral Reviews, 72, 111–128. 10.1016/j.neubiorev.2016.11.014

Morawetz, C., Hajrić, M., Rammensee, R. A., Berboth, S., & Basten, U. (2025). Wired to Regulate: Brain Connectivity Predicts Emotion Regulation Capacity and Tendency. Human Brain Mapping, 46(16), e70400. 10.1002/hbm.70400

Morris, R. W., Sparks, A., Mitchell, P. B., Weickert, C. S., & Green, M. J. (2012). Lack of cortico-limbic coupling in bipolar disorder and schizophrenia during emotion regulation. Translational Psychiatry, 2(3), e90–e90. 10.1038/tp.2012.16

Nock, M. K. (2009). Why Do People Hurt Themselves?: New Insights Into the Nature and Functions of Self-Injury. Current Directions in Psychological Science, 18(2), 78–83. 10.1111/j.1467-8721.2009.01613.x

Nock, M. K. (2010). Self-Injury. Annual Review of Clinical Psychology, 6(1), 339–363. 10.1146/annurev.clinpsy.121208.131258

Ochsner, K. N., Silvers, J. A., & Buhle, J. T. (2012). Functional imaging studies of emotion regulation: A synthetic review and evolving model of the cognitive control of emotion. Annals of the New York Academy of Sciences, 1251(1). 10.1111/j.1749-6632.2012.06751.x

Paus, T., Keshavan, M., & Giedd, J. N. (2008). Why do many psychiatric disorders emerge during adolescence? Nature Reviews Neuroscience, 9(12), Article 12. 10.1038/nrn2513

Perini, I., Gustafsson, P. A., Hamilton, J. P., Kämpe, R., Mayo, L. M., Heilig, M., & Zetterqvist, M. (2019). Brain-based Classification of Negative Social Bias in Adolescents With Nonsuicidal Self-injury: Findings From Simulated Online Social Interaction. EClinicalMedicine, 13, 81–90. 10.1016/j.eclinm.2019.06.016

Qu, D., Wen, X., Liu, B., Zhang, Xuan, He, Y., Chen, D., Duan, X., Yu, J., Liu, D., Zhang, Xiaoqian, Ou, J., Zhou, J., Cui, Z., An, J., Wang, Y., Zhou, X., Yuan, T., Tang, J., Yue, W., & Chen, R. (2023). Non-suicidal self-injury in Chinese population: A scoping review of prevalence, method, risk factors and preventive interventions. The Lancet Regional Health - Western Pacific, 100794. 10.1016/j.lanwpc.2023.100794

Quevedo, K., Martin, J., Scott, H., Smyda, G., & Pfeifer, J. H. (2016). The neurobiology of self-knowledge in depressed and self-injurious youth. Psychiatry Research: Neuroimaging, 254, 145–155. 10.1016/j.pscychresns.2016.06.015

Reinecke, A., Filippini, N., Berna, C., Western, D. G., Hanson, B., Cooper, M. J., Taggart, P., & Harmer, C. J. (2015). Effective emotion regulation strategies improve fMRI and ECG markers of psychopathology in panic disorder: Implications for psychological treatment action. Translational Psychiatry, 5(11), e673–e673. 10.1038/tp.2015.160

Robinson, K., Garisch, J. A., Kingi, T., Brocklesby, M., O’Connell, A., Langlands, R. L., Russell, L., & Wilson, M. S. (2019). Reciprocal Risk: The Longitudinal Relationship between Emotion Regulation and Non-suicidal Self-Injury in Adolescents. Journal of Abnormal Child Psychology, 47(2), 325–332. 10.1007/s10802-018-0450-6

Rolls, E. T., Cheng, W., & Feng, J. (2020). The orbitofrontal cortex: Reward, emotion and depression. Brain Communications, 2(2), fcaa196. 10.1093/braincomms/fcaa196

Silvers, J. A., Wager, T. D., Weber, J., & Ochsner, K. N. (2015). The neural bases of uninstructed negative emotion modulation. Social Cognitive and Affective Neuroscience, 10(1), 10–18. 10.1093/scan/nsu016

Sloan, E., Hall, K., Moulding, R., Bryce, S., Mildred, H., & Staiger, P. K. (2017). Emotion regulation as a transdiagnostic treatment construct across anxiety, depression, substance, eating and borderline personality disorders: A systematic review. Clinical Psychology Review, 57, 141–163.

Smoski, M. J., Keng, S.-L., Ji, J. L., Moore, T., Minkel, J., & Dichter, G. S. (2015). Neural indicators of emotion regulation via acceptance vs reappraisal in remitted major depressive disorder. Social Cognitive and Affective Neuroscience, 10(9), 1187–1194. 10.1093/scan/nsv003

Sonkusare, S., Breakspear, M., & Guo, C. (2019). Naturalistic Stimuli in Neuroscience: Critically Acclaimed. Trends in Cognitive Sciences, 23(8), 699–714. 10.1016/j.tics.2019.05.004

Steward, T., Martínez-Zalacaín, I., Mestre-Bach, G., Sánchez, I., Riesco, N., Jiménez-Murcia, S., Fernández-Formoso, J. A., Heras, M. V. de las, Custal, N., Menchón, J. M., Soriano-Mas, C., & Fernandez-Aranda, F. (2022). Dorsolateral prefrontal cortex and amygdala function during cognitive reappraisal predicts weight restoration and emotion regulation impairment in anorexia nervosa. Psychological Medicine, 52(5), 844–852. 10.1017/S0033291720002457

Swannell, S. V., Martin, G. E., Page, A., Hasking, P., & St John, N. J. (2014). Prevalence of Nonsuicidal Self=Injury in Nonclinical Samples: Systematic Review, Meta=Analysis and Meta=Regression. Suicide and Life-Threatening Behavior, 44(3), 273–303. 10.1111/sltb.12070

Tsang, M. H. L., Chen, J. C., Jiang, H., & Becker, B. (2025). Enhancing stress regulation in ecologically valid contexts through functional near-infrared spectroscopy neurofeedback of the prefrontal cortex (p. 2025.07.29.25332351). medRxiv. 10.1101/2025.07.29.25332351

Wang, S., Wu, Y., Wang, W., Zhang, J., Geng, F., Li, Q., Xiao, G., Zu, M., Nie, J., Ni, L., Zhang, D., Cheng, K., Qin, R., Ji, G.-J., & Tian, Y. (2025). Neural circuit for non-suicidal self-injury and causal clinical validation. Behavioural Brain Research, 494, 115736. 10.1016/j.bbr.2025.115736

Wang, Z., Chen, Y., Tao, Z., Yang, M., Li, D., Jiang, L., & Zhang, W. (2024). Quantifying the Importance of Non-Suicidal Self-Injury Characteristics in Predicting Different Clinical Outcomes: Using Random Forest Model. Journal of Youth and Adolescence, 53(7), 1615–1629. 10.1007/s10964-023-01926-z

Wester, K., Trepal, H., & King, K. (2018). Nonsuicidal Self=Injury: Increased Prevalence in Engagement. Suicide and Life-Threatening Behavior, 48(6), 690–698. 10.1111/sltb.12389

Whitfield-Gabrieli, S., & Nieto-Castanon, A. (2012). Conn: A functional connectivity toolbox for correlated and anticorrelated brain networks. Brain Connectivity, 2(3), 125–141.

Wilkinson, P. O., Qiu, T., Neufeld, S., Jones, P. B., & Goodyer, I. M. (2018). Sporadic and recurrent non-suicidal self-injury before age 14 and incident onset of psychiatric disorders by 17 years: Prospective cohort study. The British Journal of Psychiatry, 212(4), 222–226. 10.1192/bjp.2017.45

Wolff, J. C., Thompson, E., Thomas, S. A., Nesi, J., Bettis, A. H., Ransford, B., Scopelliti, K., Frazier, E. A., & Liu, R. T. (2019). Emotion dysregulation and non-suicidal self-injury: A systematic review and meta-analysis. European Psychiatry, 59, 25–36. 10.1016/j.eurpsy.2019.03.004

Wu, B., Zhang, H., Chen, Jinghong, Chen, Jiaye, Liu, Z., Cheng, Y., Yuan, T., & Peng, D. (2023). Potential mechanisms of non-suicidal self-injury (NSSI) in major depressive disorder: A systematic review. General Psychiatry, 36(4), e100946. 10.1136/gpsych-2022-100946

Wu, D., Wang, S., Wang, H., Zhang, J., & Tian, Y. (2025). Neural mechanisms and neuromodulation therapies for non-suicidal self-injury. Molecular Psychiatry, 30(11), 1–10. 10.1038/s41380-025-03125-7

Xiao, Q., Song, X., Huang, L., Hou, D., & Huang, X. (2022). Global prevalence and characteristics of non-suicidal self-injury between 2010 and 2021 among a non-clinical sample of adolescents: A meta-analysis. Frontiers in Psychiatry, 13. 10.3389/fpsyt.2022.912441

Yeo, B. T., Krienen, F. M., Sepulcre, J., Sabuncu, M. R., Lashkari, D., Hollinshead, M., Roffman, J. L., Smoller, J. W., Zöllei, L., & Polimeni, J. R. (2011). The organization of the human cerebral cortex estimated by intrinsic functional connectivity. Journal of Neurophysiology, 106(3), 1125–1165. 10.1152/jn.00338.2011

Zhang, L., Opmeer, E. M., van der Meer, L., Aleman, A., Ćurčić-Blake, B., & Ruhé, H. G. (2018). Altered frontal-amygdala effective connectivity during effortful emotion regulation in bipolar disorder. Bipolar Disorders, 20(4), 349–358. 10.1111/bdi.12611

Zhang, R., Gan, X., Xu, T., Yu, F., Wang, L., Song, X., Jiao, G., Liu, X., Zhou, F., & Becker, B. (2025). A neurofunctional signature of affective arousal generalizes across valence domains and distinguishes subjective experience from autonomic reactivity. Nature Communications, 16(1), 6492. 10.1038/s41467-025-61706-0

Zheng, Q., Zhou, H., Li, K., Liu, Y., Nan, W., & Gong, J. (2024). The effectiveness of mindfulness-based intervention for psychological distress and emotion regulation in college students with non-suicidal self-injury. Applied Psychology: Health and Well-Being, 16(4), 2083–2098. 10.1111/aphw.12580

Zhou, F., & Becker, B. (2025). Understanding human brain function in real-world environments. PLOS Biology, 23(6), e3003210. 10.1371/journal.pbio.3003210

Zilverstand, A., Parvaz, M. A., & Goldstein, R. Z. (2017). Neuroimaging cognitive reappraisal in clinical populations to define neural targets for enhancing emotion regulation. A systematic review. Neuroimage, 151, 105–116. 10.1016/j.neuroimage.2016.06.009

Zimmermann, K., Walz, C., Derckx, R. T., Kendrick, K. M., Weber, B., Dore, B., Ochsner, K. N., Hurlemann, R., & Becker, B. (2017). Emotion regulation deficits in regular marijuana users. Human Brain Mapping, 38(8), 4270–4279. 10.1002/hbm.23671

Zimmermann, K., Yao, S., Heinz, M., Zhou, F., Dau, W., Banger, M., Weber, B., Hurlemann, R., & Becker, B. (2018). Altered orbitofrontal activity and dorsal striatal connectivity during emotion processing in dependent marijuana users after 28 days of abstinence. Psychopharmacology, 235(3), 849–859. 10.1007/s00213-017-4803-6

