## Supplementary Information for "Naturalistic acceptance-based emotion regulation in adolescents with NSSI: altered prefrontal activation and amygdala–prefrontal connectivity"

**This file includes:**

**MRI data acquisition and preprocessing**

High-resolution structural images were obtained using Siemens' original 3D Single-shot TurboFLASH T1-weighted sequence (TR=2300 ms, TE=2.32 ms, FOV=240 mm, flip angle=8°, matrix=256×256, voxel size=0.9×0.9×0.9 mm, 192 sagittal slices, posterior-to-anterior phase encoding). Functional data were collected with an echo planar imaging–free induction decay sequence (TR=2000 ms, TE=29 ms, FOV=240 mm, flip angle=90°, matrix=80×80, voxel size=3×3×3 mm, 36 interleaved ascending axial slices, posterior-to-anterior phase encoding).

Preprocessing was conducted using Statistical Parametric Mapping (SPM12). The first five functional volumes were discarded, followed by slice-timing correction and realignment to the initial image. Head-motion–related nonlinear distortions were corrected through unwarping. Structural images were segmented and co-registered to the functional images to create skull-stripped anatomical images. Functional images were normalized into MNI space, resampled to 2×2×2 mm voxel size, smoothed with an 8-mm full-width at half maximum (FWHM) Gaussian kernel, and bias-field corrected (Gan et al., 2024; Jiang, He, Zhou, et al., 2026; Jiang, He, Zimmermann, et al., 2026). Outlier time points were detected according to the following thresholds: (a) global signal intensity more than 3 SD from the mean, or (b) signal and Mahalanobis distance values exceeding 10 mean absolute deviations based on moving averages with an FWHM of 20 volumes. All identified outlier volumes were modeled as separate nuisance regressors in the first-level analysis (Gan et al., 2024; Jiang, He, Zhou, et al., 2026; Jiang, He, Zimmermann, et al., 2026). The mean number of outlier volumes in the final sample did not differ significantly between groups (HC: 12.41±19.13, M±SD; NSSI: 11.83±15.90; $t$=0.117, $p$=0.908).

### Supplementary Tables

**Table S1.** Complement the results of subjective experience and regulation of negative emotions by Bayesian ANOVA

| Effect | $BF_{10}$ | Interpretation |
|---|---|---|
| 2 (stimulus type: NeutR, NegR)×2 (group: NSSI, HC) | | |
| StimulusType | $1.391\times10^{23}$ | Strong evidence for the alternative model |
| Group | 0.403 | Evidence favors null |
| StimulusType×Group | 1.656 | Anecdotal evidence for the alternative |
| $NeutV_{NSSI}$>$NeutV_{HC}$ | $3.144\times10^{1}$ | Strong evidence for the alternative |
| 2 (strategy: NegR, NA) × 2 (group: NSSI, HC) | | |
| Strategy | 0.147 | Evidence favors null |
| Group | 0.405 | Evidence favors null |
| StimulusType×Group | 0.181 | Evidence favors null |

NeutR, neutral video clips–react; NegR, negative video clips–react; NegA, negative video clips–acceptance; NSSI, non-suicidal self-injury; HC, health control.

### Supplementary Figures

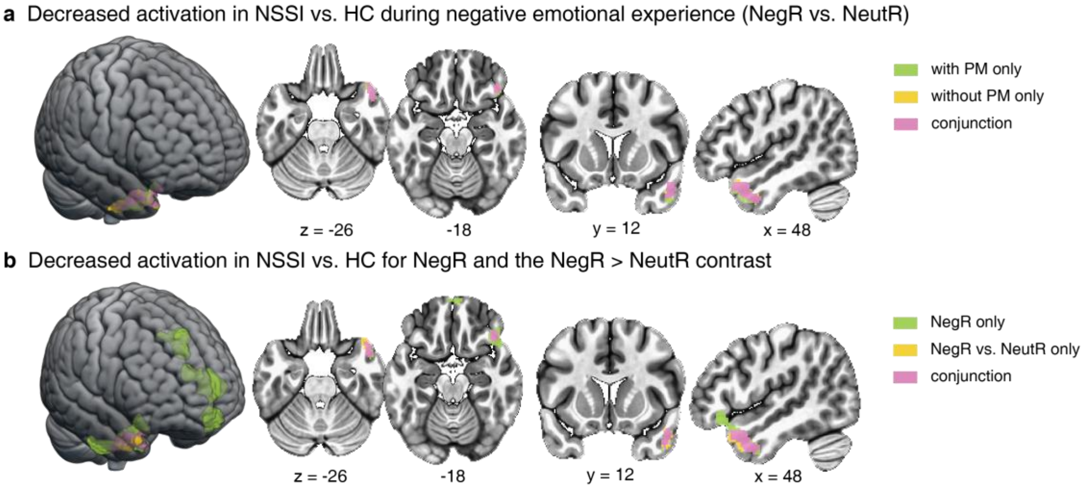

**Fig.S1 | Neural activations for negative vs. neutral reactivity and their modulation by subjective negativity. a**, Activations with (green) and without (yellow) subjective negativity ratings as parametric modulators (PM), and their conjunction (pink). **b**, Activations are shown for NegR alone (green), NegR > NeutR (yellow), and their conjunction (pink). Statistical maps are thresholded at a voxel-level $p<0.001$ and cluster-level FDR-corrected $p<0.05$. NegR, negative-view; NeutR, neutral-view. NSSI, non-suicidal self-injury; HC, health control; NeutR, neutral video clips–react; NegR, negative video clips–react; NegA, negative video clips–acceptance.

**References**

Gan, X., Zhou, F., Xu, T., Liu, X., Zhang, R., Zheng, Z., Yang, X., Zhou, X., Yu, F., Li, J., Cui, R., Wang, L., Yuan, J., Yao, D., & Becker, B. (2024). A neurofunctional signature of subjective disgust generalizes to oral distaste and socio-moral contexts. *Nature Human Behaviour*. https://doi.org/10.1038/s41562-024-01868-x

Jiang, H., He, J., Zhou, B., Guo, Y., Gan, X., Fan, X., Wang, X., Ferraro, S., Vatansever, D., Kendrick, K. M., Li, L., & Becker, B. (2026). Adolescents with non-suicidal self-injury exhibit increased pain empathic neural reactivity and personal distress to physical but not affective pain. *Journal of Affective Disorders*, *399*, 121145. https://doi.org/10.1016/j.jad.2025.121145

Jiang, H., He, J., Zimmermann, K., Zhou, X., Gan, X., Ferraro, S., Wang, L., Zhou, B., Li, L., Kendrick, K. M., Zhao, W., Yao, D., Yuan, T., Zhou, F., & Becker, B. (2026). Common and distinct neurofunctional signatures of dynamic naturalistic emotion regulation strategies. *Nature Communications*. https://doi.org/10.1038/s41467-026-70708-5
